# High seroprevalence after the second wave of SARS-COV2 respiratory infection in a small settlement on the northern coastal of Peru

**DOI:** 10.1101/2023.07.19.23292491

**Authors:** Angie K. Toledo, Franco León-Jimenez, Sofia Cavalcanti, Percy Vilchez-Barreto, Narcisa Reto, Jessica Vega, Lucia M Bolivar, Matilde Rhor, Jhon Ypanaque, Henry Silva, Luz M. Moyano, Group of Neuroepidemiology and Science of Life of Peru

**Affiliations:** School of Public Health and Administration, Universidad Peruana Cayetano Heredia, Lima, Peru; Centro de investigación para la preservación de la vida, Lima, Peru; Escuela de Medicina Humana, Universidad Cesar vallejo; Dirección de Investigación Hospital Amistad Perú-Korea Sta. Rosa II-2, Piura Peru; Centro de Salud Global, Universidad Peruana Cayetano Heredia, Tumbes, Peru; Hospital I Carlos Alberto Cortés Jiménez Essalud, Tumbes, Peru; Dirección ejecutiva de epidemiología (Chief), Centro de Salud Pampa Grande, Regional Directorate of Health, Tumbes, Peru; Centro de Salud Pampa Grande, Regional Directorate of Health, Tumbes, Peru; Psycology School (MR), Social and Cultural Office (JY), and Economy and Political Science School (HS), Universidad Nacional de Tumbes, Tumbes, Peru; Unidad Médico legal II Tumbes, Instituto de Medicina Legal del Peru.

## Abstract

**Background:** Due to more infections from variations that could escape vaccination and immunity by asymptomatic to uninfected transmission, COVID-19’s second wave had higher seroprevalence globally. Public health constraints and herd immunity may not work against these novel variations’ infectivity. This population-based study in Peru’s Tumbes Region during the second wave of COVID-19 seeks to determine seroprevalence and demographic changes from the first wave.

**Methodology/Principal findings:** In Dec 2021-Jan 2022, a study in Tumbes’ informal settlement sampled individuals over 2 years old from one in every four households. Finger-prick blood samples and symptom surveys were collected. On the second wave, there was a substantial rise in adjusted seroprevalence (50.15%, 95% CI [45.92 – 54.40]) compared with the first wave (24.82 %, 95%CI [22.49 – 27.25]), with females maintaining a higher seroprevalence (53.89; 95% CI [48.48-59.23]) vs. 45.49; 95% CI [38.98-52.12], p=0.042) compare to males. Those under 18 years of age had the highest IgG seropositivity: the 12–17 age group during the second wave (85.14%) and the 2–11 age group (25.25%) during the first wave. Nasal congestion and cough were symptoms associated with seropositivity, unlike the first wave.

**Conclusions/Significance:** In Tumbes, the seroprevalence of COVID-19 increased by twofold compared to the initial wave. Inadequate infrastructure and limitations in human resources and supplies in healthcare facilities made the Peruvian health system collapse. We must include in epidemiological surveillance mHealth tools that enable real-time reporting of new cases. Working alongside the community is the only way to improve any new intervention strategy to prevent or control a new pandemic.

**Author summary:** In Peru, the healthcare system was overwhelmed by the COVID-19 pandemic due to the lack of hospital capacity, oxygen supply, political unrest, and a fragmented healthcare system. During the first wave, the prevalence ranged from 20.8% to 72%, and it was predicted that the second wave would be disastrous. To assess the seroprevalence of SARS-CoV-2, a cross-sectional study was conducted in the settlement “AAHH Las Flores” located in front of Tumbes National University’s main campus. A door-to-door intervention was conducted, and a total of 580/781 (74.26%) individuals over than 2 years and above agreed to participate. After adjusting for sensitivity and specificity, the calculated adjusted seroprevalence was 50.15%. Women had a slightly higher adjusted seroprevalence compared to men, and the age groups with the highest prevalence of IgG seropositive were from 12 to 17 years, from 30 to 59 years, and older than 60 years. More than 80% of seropositive patients were asymptomatic in all age groups. The study’s findings suggest that COVID-19 transmission in the settlement was higher during the second wave, and asymptomatic individuals could have played a critical role in spreading the virus.

## Introduction

Due to a lack of hospital capacity, critical oxygen supply, political unrest, and a fragmented healthcare system, Peru experienced high rates of morbidity and mortality during the first and second catastrophic waves (1–3). Prevalence rates on the first wave ranged from 20.8% to 72% (1,2,4,5). Without the option of massive vaccination, a disastrous second wave was expected unless the health system responded adequately. On December 17th, 2021, in the Tumbes district, close to 93% of the population over 18 years old and seniors had received the first dose of the Sinopharm and Pfizer COVID-19 vaccine. 75% of the population between the ages of 12 and 17 was vaccinated, while no one under the age of 12 had been immunized (6). The first wave hit Peru in March 2020, peaked in August, and declined in December 2020 (7). The second wave started in January, with the highest number of infections and deaths decreasing in June 2021. Throughout a significant portion of the second wave, the number of daily infections consistently exceeded the number of daily recoveries. The peak was observed during week 54, specifically from March 22nd to 28th, with an average of 9,079 new infections and 8,727 recoveries per day (8,9). The highest attack rate (6.96) was for 30-to 59-year-olds, followed by 6.56 for adults aged 60 and above. (9). During this same period, Tumbes had the seventh highest cumulative attack rate (5.29), compared with the national average of 4.84. The Tumbes case fatality rate rose from 3.54% to 9.40% on Maýs last day of 2021. Peru was fifth in the world for deaths per 100,000 people (10), behind the United States, Brazil, India, and Mexico (2,9). In April 2021, during the peak of cases, the Tumbes region had three hospitals, 17 beds in the ICU, and 20 mechanical ventilators(11).

A cross-sectional study conducted in a rural village of Tumbes (Puerto Pizarro) during the final days of the first wave found an adjusted seroprevalence of 24.82% (12) where; with being a woman [28.03%, p=0.002], water storage [PR 1.37, p=0.034], and symptoms such as fever, general discomfort, cough, nasal congestion, respiratory distress, headache, anosmia, and ageusia being variables associated with a positive antibody rapid-test of Sars-cov-2 (12,13). Taking full advantage of the regional program of active surveillance of the Tumbes Government (GORE) and Regional Directorate of Health (DIRESA) (14), we conducted a cross-sectional study in a human settlement named AAHH Las Flores (n = 781 population): a) to assess seroprevalence of SARS-CoV-2 at the end of the second wave, b) to determine distribution by age group, and health determinants associated with residents over 2 years old.

## Materials and methods

### Ethical considerations

Approval for the study protocol and consent forms was obtained from the institutional review boards of Universidad Peruana Cayetano Heredia and the Regional Directorate of Health (DIRESA, Spanish acronym) in Tumbes. To ensure comprehension, illiterate individuals were included by having the IC and survey read aloud to them. All participants provided written informed consent (IC) in the presence of a witness (S1 Text). For minors, both the minor and their parents or legal guardians provided written informed consent (S2 Text).

### Area and study population

The settlement “AAHH Las Flores,” which has a population of 781 individuals, is located in the Pampa Grande Population Center in the district and province of Tumbes. It is situated 2.5km away from the town center and lacks proper infrastructure, with unpaved streets and no continuous access to safe drinking water (15). The population is of a homogeneous mestizo ethnicity. According to the last national census, the population of this settlement was 721 individuals, with a total of 184 households. Also, 94.5%, had access to potable water and 95.1% had access to sewage disposal (16).

The Pampa Grande health facility was staffed by six physicians, six nurses, six midwives, one pediatrician, and six nursing technicians. It provides 24/7 service and has an ambulance for emergency transportation (17).

### Study design and sampling

A cross-sectional analytical approach was used to invite and enroll individuals over than 2 years and above (n=781 pop) through a door-to-door intervention conducted between December 2021 and the first day of January 2022. This intervention took place after the second wave and a few weeks before the start of the third wave.

The recruitment process, censuses, and rapid antibody testing for COVID-19, were described elsewhere (12,13). Briefly, the regional directorate of Health-Tumbes and the Center for Global Health collaborated to conduct a nominal census with block-numbered homes. Individuals over the age of two who stayed in the village at least three nights per week and consented to sign the informed consent form were eligible to participate. The study included 781 individuals who met these criteria. Non-medical field workers collected blood samples via finger prick and explained the results of the lateral flow test to everyone surveyed. Clinical signs suggestive of COVID-19 disease were evaluated in accordance with WHO criteria (18). Trained physicians from the study team evaluated participants with respiratory symptoms, and data was entered into the Notiweb and SISCOVID databases for follow-up by the healthcare system. The COVID-19 Clinical Epidemiological Investigation format was used to identify symptoms experienced within the previous 14 days, as recommended by the Peruvian Ministry of Health (19).

A census previously validated by the Center for Global Health in epidemiological population-based investigations was used in this study (20,21). Comorbidities were determined as the existence of at least one of the following medical disorders: hypertension, diabetes, hepatic disease, renal disease, lung disease, asthma, obesity, neurological problems, HIV, cancer, or tuberculosis (22).

### Definitions and operationalization of variables

The seroprevalence of SARS-CoV-2 was determined by dividing the number of participants who tested positive for IgG, IgM, or both in a lateral flow test by the total number of participants. The variables: sex, “type of family”, rooms per person, water supply and water storage were describe in a previously research (12). “Latrines/don’t have” and “other and restrooms,” which are designated as places with a toilet, sink, network connection, and drainage, are the two categories for the variable for hygienic services (12). Visual examination was used to confirm the factors related to the water supply, water storage, and hygiene services. A lateral flow test was declared positive when IgG, IgM, or both findings were observed.

### Statistical analysis

The SARS-CoV-2 seroprevalence ratios in Las Flores settlement were aimed to be estimated by performing descriptive statistics and binomial family generalized linear models with a logarithmic link function, while controlling for other variables, such as age, sex, type of family, water storage, and basic hygienic services at home. Household clustering was accounted for by using robust sandwich-type standard errors. Each variable was evaluated for inclusion in the final model using the log likelihood ratio. 95% confidence intervals (CIs) were reported, and statistical significance was set at P < 0.05. The seroprevalence was adjusted for the test’s reported sensitivity (99.03%) and specificity (98.65%). The statistical analysis was performed using Stata v 17.0 software (College Station, Texas 77845, USA).

## Results

A total of 580/781 (74.26%) individuals older than 2 years old agreed to participate. Female participants were 59.83 % (347/580). The overall participants’ mean age was 28.80±19.36 years. MINSA stratification mean ages were from: 2 to 11 years (mean 6.83±2.81), 12 to 17 years (mean 14.53±1.81), 18 to 29 years (mean 23.60±3.23), 30 to 59 years (mean 44.00±7.98) and older than 60 years (mean 67.84±6.80). Most people lived in single families 391/580 (67.41 %), had access to public potable water 342/580 (58.97%) for 02-04 hours per day and 454/580 (78.28 %) had access to hygienic services.

### Seroprevalence

Anti-SARS-CoV-2 antibodies were detected in 292/580 (50.34 %) participants: 282 IgG reactive, 8 IgM/IgG reactive, and 2 IgM reactive. After adjusting for sensitivity (99.03%) and specificity (98.65%), the calculated adjusted seroprevalence was 50.15 % (95% CI: 45.92 - 54.40). Women had a slightly higher adjusted seroprevalence compared to men (53.89 [95% CI: 48.48 - 59.23] vs. 45.49 [95% CI: 38.98 - 52.12], p=0.042). No other demographic characteristics were associated with SARS-CoV-2 lateral flow test seropositivity (Table 1).

**Table 1.** Seroprevalence adjusted for participant characteristics and associated factors from first and second COVID-19 wave.

As illustrated in Fig 1, the age groups with the highest prevalence of IgG seropositive were from 12 to 17 years (85.14%), from 30 to 59 years (59.80 %) and older than 60 years (55.81%). On the other hand, more than 80% of seropositive patients were asymptomatic in all age groups (Figure 2).

**Fig 1.**
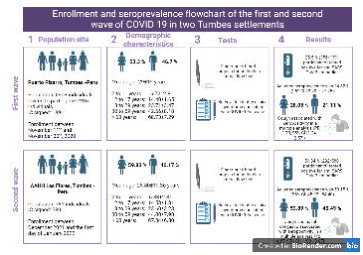
Seronegative and seropositivity from first and second COVID-19 wave.

**Fig 2.** Symptomatic and asymptomatic cases in the seropositive population from first and second COVID-19 wave.

In a bivariate analysis, symptoms such as cough (PR: 1.74; 95% CI: [1.27 - 2.38], p=0.001) and nasal congestion (PR: 0.60, 95% CI: [0.36 - 0.99], p=0.045) were associated with SARS-CoV-2 seropositivity. Both symptoms were associated with seropositivity in a multiple analysis (PR: 4.15, 95% CI: [1.90 - 9.07], p<0.001) and (PR: 0.41; 95% CI: [0.20 - 0.84], p=0.014) (Table 2).

**Table 2.** Self-reported symptoms seropositive and seronegative participants from first and second COVID-19 wave.

## Discussion

In the second wave, a higher COVID-19 prevalence was reported globally compared to the first wave. In Peru, it grew by 100 percent between November 2020 (first wave) and December 2021 (second wave). This research group conducted two population-based studies in the Tumbes Region, revealing the same trend (24.82% vs. 50.15%) (S1 Fig), contradictory to Regional directorate of health Tumbes reported (6.11%) (14). This numbers were similar to reported in Italy, which had a seroprevalence of over 50% in the second wave (23), and higher than Japan, which doubled its caseload (24). Seroprevalence increased as more people were infected with the virus and its different variants that could escape vaccination (25) and the immunity generated by contagion from the asymptomatic to the uninfected (26). In addition, these variants resulted in variations in transmissibility, disease severity, risk of reinfection, diagnostic failures, natural immunity, vaccination efficacy, and treatment. Persistent community transmission in regions with a higher vaccination rate or history of infection may indicate the presence of a variant capable of evading the immune response (27). The new variants began to appear in the world at the end of 2020, right at the end of the first study, as variants of interest (VOI) and variants of concern (VOC) according to the WHO; the Delta variant began to circulate in October 2021 and could be the variant that explains this high seroprevalence (28). According to a weekly report from the INS, in Peru, at the end of August 2021, the delta variant corresponded to 40% of the samples, gamma was 37%, lambda was 17%, and the mu variant was 5% (29). In Tumbes, in the 20–30-year-old group. From January to November 2022, omicron (n = 463) and delta (n = 125) variants were identified (30).

The delta variant generated a lower response to mRNA vaccines (Pfizer-BioNTech and Moderna NIH), viral vectors (Oxford-AstraZeneca) (31), as well as inactivated viruses (Sinopharm-Beijing) (32). On December 19, 2022, the coverage with three doses of vaccines in Tumbes was as follows: 55.1% for adolescents, 77.15 % for young persons, 77.9 % for adults, and 82.5 % for the elderly. Indicative of adequate coverage, the majority of doses were distributed by Pfizer (56%), AstraZeneca (19.7%), Sinopharm (17%), and Moderna (7.2%) (33). In addition, the WHO classified the omicron variant as concerning on November 26, 2021. Although the classic public health restriction measures may be effective against the new variants, they are less effective in terms of infectivity; on the other hand, herd immunity achieved with a previous variant might not be helpful (28).

Seroprevalence in the second wave (53.89 [48.48-59.23]) was higher compared to other Reports of the second wave in Peru, where there were no gender differences (2,14), similar in France (Corsica) (34); our numbers were lower compared to the Democratic Republic of the Congo (74.3% vs. 68.3%; p = 0.021) (35), Kashmir, India (86.21 [85.86-86.55](36) and slightly higher compared to Delhi, India (52.42,OR 1.16[1.10-1.23],p<0.001)(37). A possible explanation for this increased seroprevalence in women is that they were the ones caring for positive cases at home (38); there are more female health workers than male health workers (nurses, nurse technicians, and midwives) (39); women purchase supplies in stores and markets, and this overexposure could lead to an increase in COVID-19 infection.

There were many possible explanations for the only statistically significant clinical manifestations in this second wave (cough and nasal congestion): the new variants sacrificed their virulence for their infectious capacity, so we observed in the delta variant there was some variability in clinical expression compared to the original virus strain (40); people gained more knowledge about COVID-19 on the second wave, may have contributed to the increased prevalence of clinical manifestations.

One of the limitations of this research is the sample size and the fact that the study was conducted in a single settlement, limiting the generalizability of the findings to other regions of Peru. The limited availability of rapid tests, which have lower sensitivity and specificity compared to PCR tests; and finally, the study did not collect data on participants’ vaccination status; this information was obtained from reports of the Ministry of Health. With persistent IgG seropositivity, it is recommended that follow-up be conducted to detect COVID-19 sequelae related to the presence of pulmonary lesions, mental health issues and, the implementation of an application-based surveillance system in our country. Many systematic reviews and meta-analyses revealed that 62.2% of asymptomatic patients had tomographic abnormalities, indicating that they are a potential source of transmission (41)

Our research has several implications for Peru’s public health policy. First, the high seroprevalence indicates that a substantial portion of the population may already be infected, indicating the need to increase vaccine coverage. The significance of targeted vaccination campaigns to safeguard younger age groups with a higher seroprevalence is emphasized by the study’s findings. Given the higher seroprevalence in females, there may be gender-specific differences in exposure to the virus that need further investigation.

## Conclusions

This study shows a higher prevalence of SARS-CoV-2 antibodies after the second wave of COVID-19 in Tumbes, with a huge prevalence of IgG in individuals younger than 17 years old, more than 80% of seropositive patients were asymptomatic in all age groups. The findings suggest that a significant portion of the population was exposed to the virus and developed antibodies, highlighting the importance of continued efforts to control the spread of new variants of the virus through massive campaigns of vaccination.

## Data Availability

All data are fully available without restriction

## Acknowledgments

We thank the villagers of AAHH Las Flores. LMM and team are grateful to all the regional authorities from the County Town Hall, the Regional Directorate of Health-Tumbes V Jimenez, and J Arias. We are grateful to the members of the Executive Office of Epidemiology N Julca, and L Arevalo. LMM expresses gratitude to the members of the Grupo de Investigación de la Escuela de Medicina. LMM would like to thanks for the aerial photography to JeanPierre. Angie Toledo is doctoral student in the Epidemiological Research Doctorate at Universidad Peruana Cayetano Heredia under FONDECYT/CIENCIACTIVA scholarship EF033-235-2015 and supported by training grant D43 TW007393 awarded by the Fogarty International Center of the US National Institutes of Health. Neither the FONDECYT/CIENCIACTIVA scholarship nor the training grant D43 TW007393 contributed to the development of the present study.

## Footnotes

The statements contained in this document are those of the authors and should not be construed as official points of view of the Universidad Cesar Vallejo, Universidad Nacional de Tumbes, or other organizations mentioned.

## Supporting information

S1 Data. Excel spreadsheet containing, in separate sheets, the underlying data used for the statistical analysis and description of variables.

S1 Text. Adult informed consent for study participation approved by the institutional review boards of Universidad Peruana Cayetano Heredia.

S2 Text. Parent-signed informed consent for minor study participation approved by the institutional review boards of Universidad Peruana Cayetano Heredia.

S1 Fig. Enrollment and seroprevalence flowchart of the first and second wave of COVID-19 in two Tumbes settlements. Created with BioRender.com

